# Joint modelling of individual trajectories, within-individual variability and a later outcome: systolic blood pressure through childhood and left ventricular mass in early adulthood

**DOI:** 10.1101/19008474

**Authors:** Richard M.A. Parker, George Leckie, Harvey Goldstein, Laura D. Howe, Jon Heron, Alun D. Hughes, David M. Phillippo, Kate Tilling

## Abstract

Within-individual variability of repeatedly-measured exposures may predict later outcomes: e.g. blood pressure (BP) variability (BPV) is an independent cardiovascular risk factor above and beyond mean BP. Since two-stage methods, known to introduce bias, are typically used to investigate such associations, we introduce a joint modelling approach, examining associations of both mean BP and BPV across childhood to left ventricular mass (indexed to height; LVMI) in early adulthood with data from the UK’s Avon Longitudinal Study of Parents and Children (ALSPAC) cohort. Using multilevel models, we allow BPV to vary between individuals (a “random effect”) as well as to depend on covariates (allowing for heteroscedasticity). We further distinguish within-clinic variability (“measurement error”) from visit-to-visit BPV. BPV was predicted to be greater at older ages, at higher bodyweights, and in females, and was positively correlated with mean BP. BPV had a positive association with LVMI (10% increase in SD(BP) was predicted to increase LVMI by mean = 0.42% (95% credible interval: −0.47%, 1.38%)), but this association became negative (mean = −1.56%, 95% credible interval: −5.01%, 0.44%)) once the effect of mean BP on LVMI was adjusted for. This joint modelling approach offers a flexible method of relating repeatedly-measured exposures to later outcomes.

## INTRODUCTION

When exposures vary over time – for example systolic blood pressure (BP) as measured over the life course – both the overall mean and the change in the exposure can affect later health outcomes (1). However, within-individual variability – i.e. the extent to which an individual’s measure fluctuates around this trend – may also be a risk factor (2).

Methodological difficulties have meant that within-individual variability in time-varying exposures have been seldom examined, and when they are it is often using two-stage procedures, known to introduce bias (3). An alternative, one-stage, approach is to use joint multilevel models to relate time-varying exposures (4) to a later outcome (5). These models have recently been extended to also examine within-individual variability (3). Such joint models leads to unbiased parameter estimates and correct standard errors: either when relating the mean and trajectory (6), or within-individual variability (3), of an exposure to a later outcome.

We illustrate this approach by examining the association of both mean BP and within-individual blood pressure variability (BPV) with an established biomarker of target organ damage: left ventricular mass indexed to height (LVMI). Whilst BPV can be measured across various time scales (7), here we focus on the longer-term, a.k.a. “visit-to-visit”. In adults, visit-to-visit BPV has been found to be an important predictor of subsequent cardiovascular disease (CVD) over and above mean level (8). Detecting those at risk of CVD early in life is important in order to design and administer preventative measures in a timely and targeted manner (9, 10). Mean systolic BP in childhood is positively associated with early signs of heart damage, such as left ventricular hypertrophy (11-13), and there is evidence from a US cohort of children that higher BPV – as estimated via the standard deviation (SD) of the measurements across all visits (conducted every 2-3 years) for each person – is associated with adult hypertension, independent of mean BP levels (14). However, there has been little investigation of the role of BPV on early target organ damage, despite its potential utility as a predictive factor (15), and only limited analysis of the factors associated with childhood BPV (14, 16).

Our aim was to explore the factors associated with within-individual BPV in a UK cohort of children using multilevel analyses, and to extend these to joint models to investigate the association of both mean BP and BPV with LVMI in early adulthood.

## METHODS

### Participants

The participants are from the Avon Longitudinal Study of Parents And Children (ALSPAC), an ongoing prospective longitudinal birth cohort study based in SW England (17-19). Pregnant women resident in Avon, UK with expected dates of delivery 4/1/1991 to 12/31/1992 were invited to take part in the study. The initial number of pregnancies enrolled was 14,541 resulting in 13,988 children alive at 1-year of age. When the oldest children were approximately 7 years of age, an attempt was made to bolster the initial sample with eligible cases who had failed to join the study originally, ultimately increasing the number of enrolled pregnancies to 15,454, with 14,901 children alive at 1-year of age. The study website (http://www.bristol.ac.uk/alspac/researchers/our-data/) contains details of all the data that is available through a fully searchable data dictionary and variable search tool.

5,217 of the participants attended an ALSPAC study clinic at mean age 17.7 years, of whom a random subsample of 2,047 (all singletons) had their LVMI (left ventricular mass in grammes, indexed to height in m^2.7^, i.e. g/m^2.7^) measured via echocardiography (20, 21) (exclusion criteria included pregnancy and congenital heart disease; see below for a comparison of the subsample modelled with the larger sample enrolled in ALSPAC, and Boyd *et al*. (17) for a discussion of attrition in ALSPAC). Echocardiography was performed using an HDI 5000 ultrasound machine (Phillips Healthcare, Amsterdam, The Netherlands) equipped with a P4-2 Phased Array ultrasound transducer by one of two experienced echocardiographers using a standard examination protocol (21).

1,988 of these participants had their systolic BP (SBP), height and weight and age recorded, on at least one prior occasion (research clinic). These clinics were at *c*. 7.5-, 9.5-, 10.5-, 11.5-, 13-, 15.5-years of age. At each clinic, the participant’s SBP (in mmHg) was measured at least twice, using a validated electronic monitoring device and a cuff size appropriate for their upper arm circumference, with the participant sitting and at rest with the arm supported. Each of these measurements was available for our analysis from each clinic, apart from the clinic at *c*.10.5 years of age, for which only the mean was available.

### Statistical analysis

The covariates we included for SBP were age (for further details, see below), sex, an interaction between age and sex, weight, height, and a number of maternal characteristics including parity (the number of previous pregnancies resulting in either livebirth or stillbirth), age at delivery, and highest educational qualification. Sex, age and weight, as measured at the clinic at *c*.17.7 years of age, were included as exposures for LVMI, but height was not, as LVMI is indexed to height. All continuous covariates were centred around their grand mean prior to analysis. Weight, as measured across childhood, and LVMI were log-transformed prior to analysis.

We used a joint model to relate BPV to subsequent log(LVMI). We describe this modelling approach below, building up model complexity using an example which assumes that SBP increases linearly with age across childhood (we later relax this assumption).

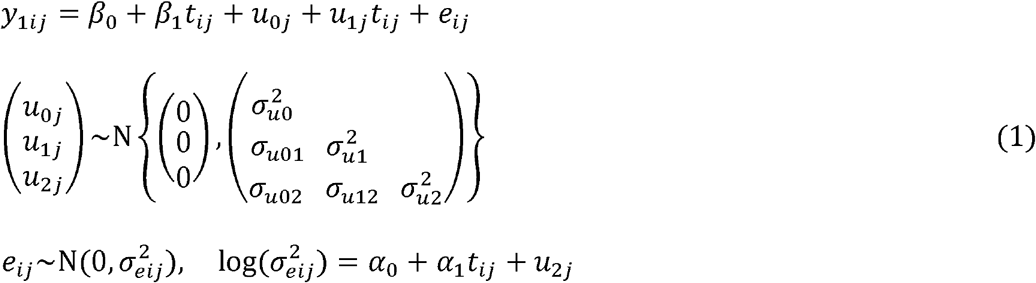

In equation 1, *y*_1*ij*_ is the repeatedly-measured exposure (SBP) measured at clinic *i* (*i* = 1, …, *n*_*j*_) for individual *j* (*j* = 1, …, J), with the covariate *t*_*ij*_ indicating the individual’s age at that clinic. In the mean function for BP, there are individual-level random effects for the intercept (*u*_0*j*_; mean BP at *t* = 0) and also for rate of change (*u*_1*j*_; BP slope), with within-individual (between-clinic) error *e*_*ij*_. In standard multilevel models, the variance of the within-individual error is assumed to be constant (homoscedastic) across all observations (as 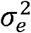). Here it is allowed to depend on age (*t*_*ij*_), and also on an individual-level random effect *u*_2*j*_ (with a log-link ensuring that the within-individual variance remains positive). Within-individual variability in blood pressure, log(BPV), is therefore allowed to change with age (as estimated here via *α*_1_). The random effect *u*_2*j*_ allows each individual to have their own estimate of log(BPV). This is a mixed-effects location scale model, as described and demonstrated by Hedeker *et al*. (22, 23); see also Goldstein *et al*. (24). It is also possible to allow for coefficients of covariates within the within-individual variability function to randomly-vary across individual: for example allowing the coefficient for age (*t*_*ij*_) to randomly-vary to investigate whether its association with BPV differs between people (25).

This model can be expanded to include the later individual-level outcome (log(LVMI), measured at *c*.17.7 years), producing a joint model with shared random effects, as follows:

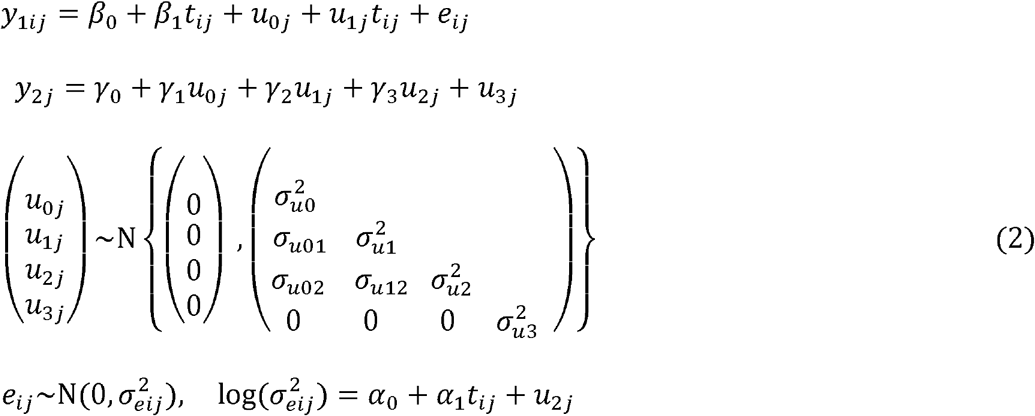

In equation 2, *y*_2*j*_ denotes log(LVMI) for individual *j* (*j* = 1, …, J). The random effects for mean BP, BP slope and log(BPV), discussed above, are included as exposures in the linear model for the mean of log(LVMI). (See Appendix 1 for an alternative parameterisation).

The models above have two levels, and assume there is just one measure per person, per clinic. If more than one measurement of SBP is taken for each individual at each clinic session, however, then the model can be expanded as follows:

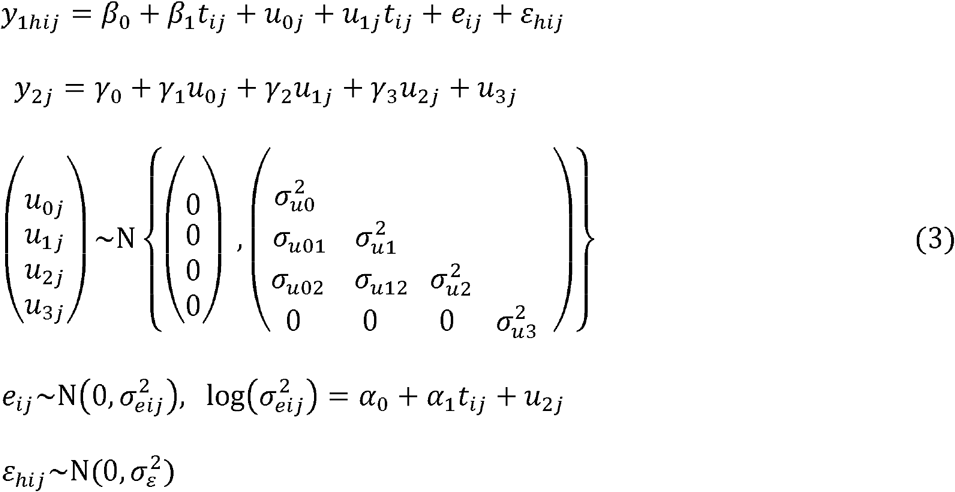

In equation 3, *y*_1*hij*_ denotes SBP as measured at (within-clinic) occasion *h* (*h* = 1, …, *n*_*ij*_) for clinic *i* (*i* = 1, …, *I*_*j*_), for individual *j* (*j* = 1, …, J). *ε*_*hij*_ refers to the residual within-clinic error in the repeatedly-measured exposure, assumed to have constant variance 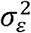. This within-clinic measurement error, which we assume is not related to log(LVMI) (*y*_2*j*_), will incorporate both systematic (e.g. white coat effect / habituation) and random error.

Following previous work modelling SBP across these ALSPAC clinics, we allowed for a non-linear relationship between age and mean SBP by fitting a linear spline with a knot point at 12 years (26, 27). The spline variables were derived as follows (where *t*_*ij*_ = age in years) (28):

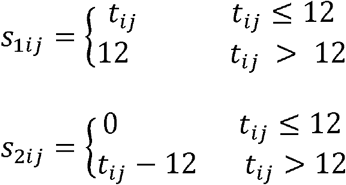

These spline terms were fitted as fixed effects in the mean function for SBP. Since allowing the coefficients of these terms to randomly-vary across individuals lead to convergence problems in some models, a linear term for age fitted across the whole age range was fitted as a random effect. We assumed that BPV had a linear association with age.

Appendix 2 includes estimates from sensitivity analyses designed to (a) investigate the influence of the submodel for log(LVMI) on the random effect estimates; (b) compare a parameterisation modelling clinic mean BP with current models; (c) check whether restricting the analysis sample to those with LVMI measures had any effect on model for change in SBP, examining possible selection bias; (d) investigate sex as the only observed covariate, for comparison with published findings elsewhere (14, 16).

We used Bayesian estimation via MCMC methods in Stan (2.19.1) (29, 30), called from R (31) using the rstan package (32). See Appendix 3 for examples. Appendix 4 provides further details of estimation.

Ethical approval for the study was obtained from the ALSPAC Ethics and Law Committee and the Local Research Ethics Committees. Informed consent for the use of data collected via questionnaires and clinics was obtained from participants following the recommendations of the ALSPAC Ethics and Law Committee at the time.

## RESULTS

Two observations at the individual-level, eight at the clinic-level, and one at the measurement-level (36 measurement-level observations in total) were identified as outliers in quantile-quantile plots of residuals from preliminary models and were removed from the dataset prior to further analyses. The resulting dataset comprised 1,986 individuals attending 10,556 clinic sessions with 19,360 recorded BP measurements.

Table 1 compares the participants included in the analysis with those 12,318 ALSPAC children who were not included (but were recruited in an eligible ALSPAC phase, singletons, and otherwise recorded as alive at 1 year of age). There was evidence that the included participants were more likely to be female, had mothers who had had fewer children and were more educated and healthier than those not included.

**Table 1.**
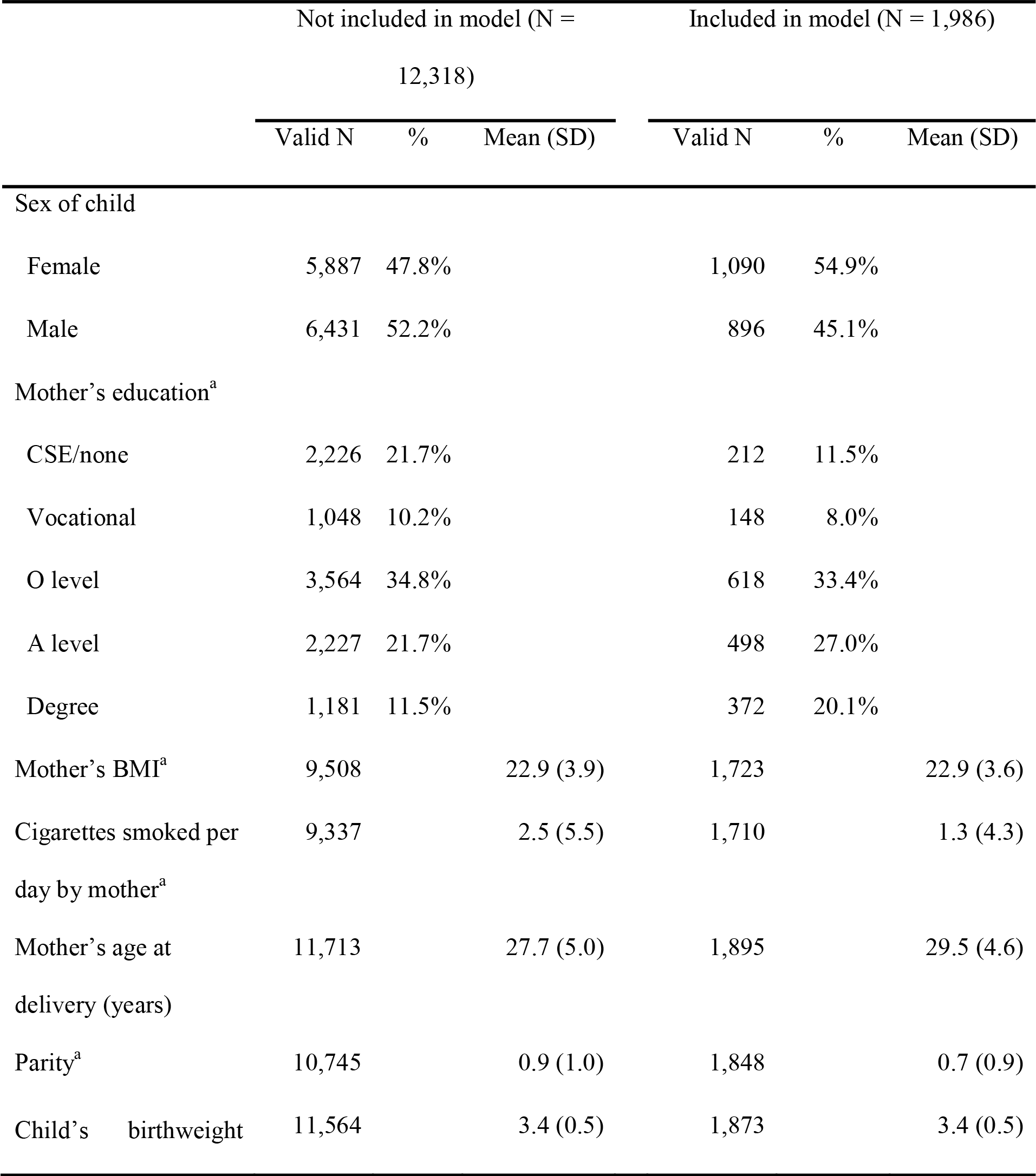
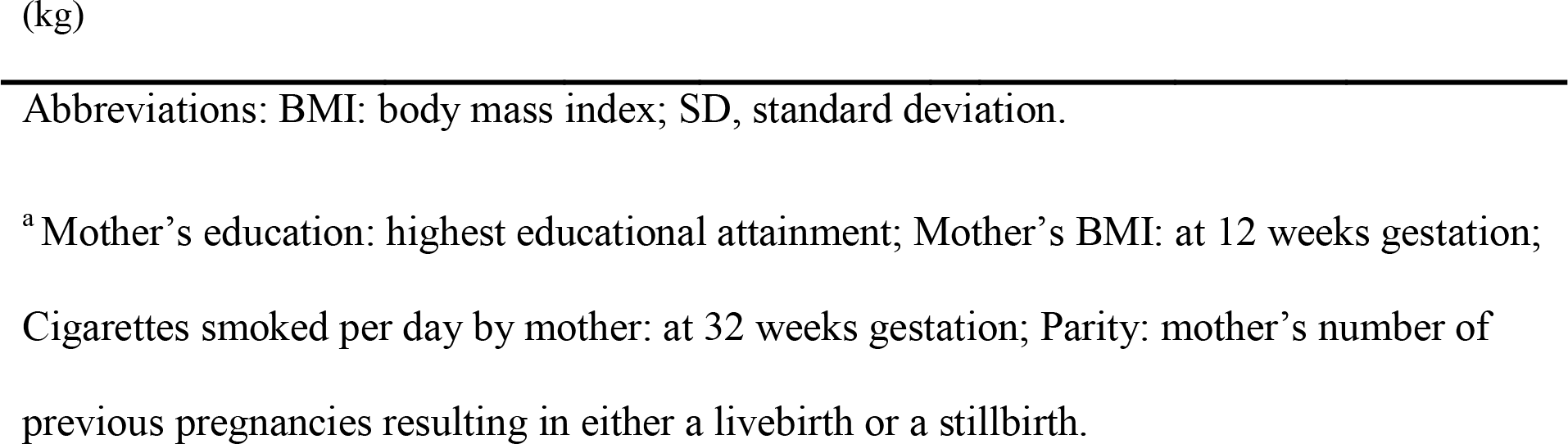
Comparing Characteristics of Children Later Undergoing Echocardiography, and Included in the Model, to Those not Included in the Model, From the Avon Longitudinal Study of Parents And Children, SW England, 1991-2008.

|  | Not included in model (N = 12,318) |  |  | Included in model (N = 1,986) |  |  |
| --- | --- | --- | --- | --- | --- | --- |
|  | Valid N | % | Mean (SD) | Valid N | % | Mean (SD) |
| Sex of child |  |  |  |  |  |  |
| Female | 5,887 | 47.8% |  | 1,090 | 54.9% |  |
| Male | 6,431 | 52.2% |  | 896 | 45.1% |  |
| Mother's education<sup>a</sup> |  |  |  |  |  |  |
| CSE/none | 2,226 | 21.7% |  | 212 | 11.5% |  |
| Vocational | 1,048 | 10.2% |  | 148 | 8.0% |  |
| O level | 3,564 | 34.8% |  | 618 | 33.4% |  |
| A level | 2,227 | 21.7% |  | 498 | 27.0% |  |
| Degree | 1,181 | 11.5% |  | 372 | 20.1% |  |
| Mother's BMI<sup>a</sup> | 9,508 |  | 22.9 (3.9) | 1,723 |  | 22.9 (3.6) |
| Cigarettes smoked per day by mother<sup>a</sup> | 9,337 |  | 2.5 (5.5) | 1,710 |  | 1.3 (4.3) |
| Mother's age at delivery (years) | 11,713 |  | 27.7 (5.0) | 1,895 |  | 29.5 (4.6) |
| Parity<sup>a</sup> | 10,745 |  | 0.9 (1.0) | 1,848 |  | 0.7 (0.9) |
| Child's birthweight | 11,564 |  | 3.4 (0.5) | 1,873 |  | 3.4 (0.5) |

Abbreviations: BMI: body mass index; SD, standard deviation.
<sup>a</sup> Mother's education: highest educational attainment; Mother's BMI: at 12 weeks gestation; Cigarettes smoked per day by mother: at 32 weeks gestation; Parity: mother's number of previous pregnancies resulting in either a livebirth or a stillbirth.

Figure 1 plots mean SBP against mean age for each clinic for those included in the model, indicating sample sizes for each clinic. The mean number of clinics attended was 5.3.

**Figure 1.**
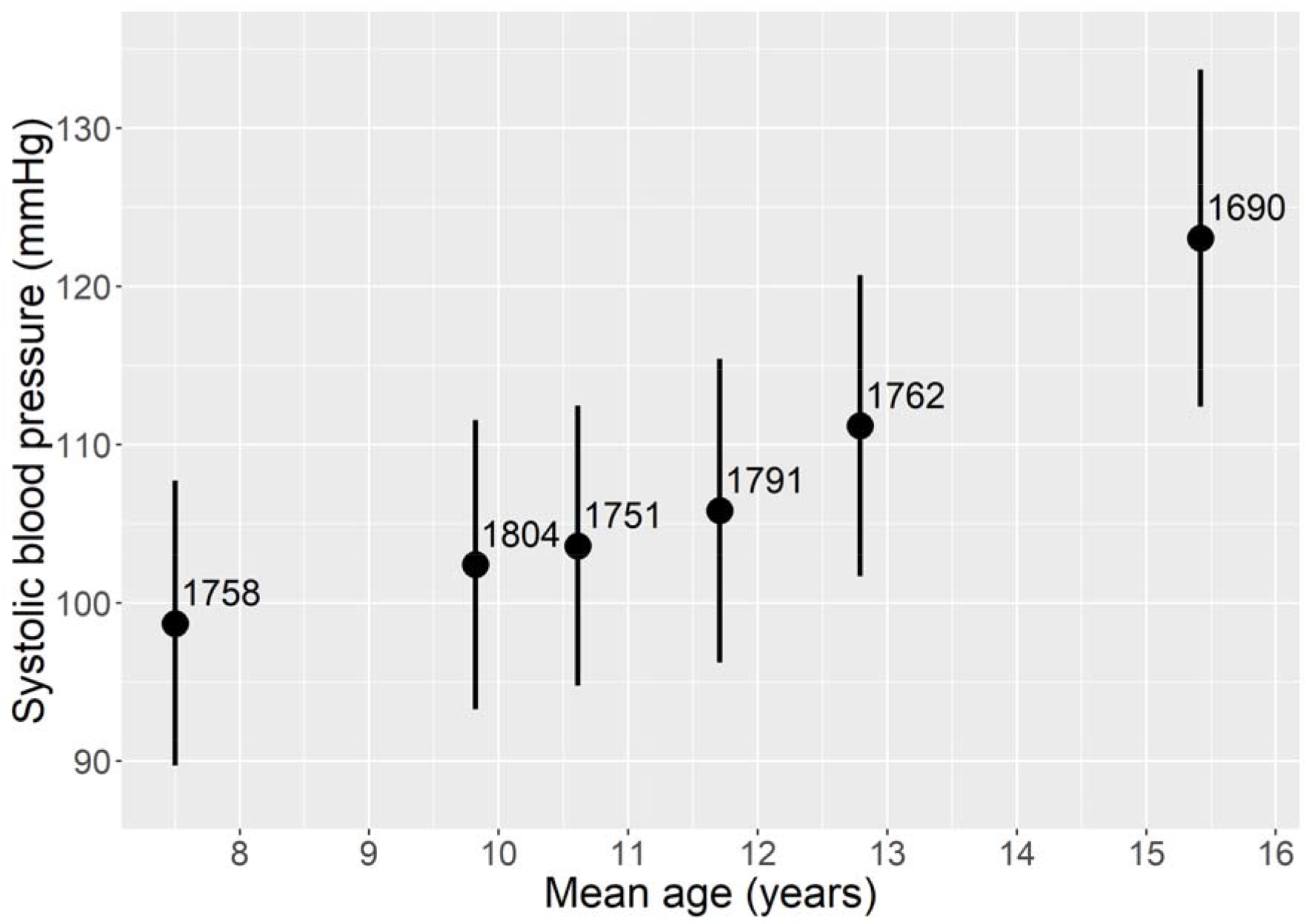
Mean clinic systolic blood pressure against mean age for clinics attended by those participants later undergoing echocardiography; within-plot annotations indicate sample size (individuals). Avon Longitudinal Study of Parents And Children, SW England, 1991-2008. Bars: 1SD either side of mean.

Table 2 summarises 3-level joint analyses in which both SBP trajectory and log(LVMI) are simultaneously modelled, with age centred around the sample mean of 11.3 years. With just age and sex as observed covariates (in model A), mean SBP was predicted to increase with age. The rate of increase was similar for males and females up to the age of 12 years, with mean BP predicted to increase more steeply for males at older ages. With regard to within-individual variance, with each year of age, BPV was predicted to increase by mean = 15.6% (95% credible interval (CI): 12.9%, 18.4%), or if expressed as the predicted increase in SD(BP): mean = 7.5% (95% CI: 6.2%, 8.8%). Females’ BPV was predicted to be mean = 16.7% (95% CI: 4.5%, 30.4%) greater than that for males (SD(BP): mean = 8.0% (95% CI: 2.2%, 14.2%)). Age at time of echocardiography was estimated to have a modest negative association with LVMI, and females were predicted to have smaller values of LVMI.

**Table 2.** Estimates From Joint Models With Shared Random Effects, Analysing SBP and log(LVMI) as Outcomes, Presenting the Posterior Parameter Estimates of the Regression Coefficients (Except Where Indicated), Avon Longitudinal Study of Parents And Children, SW England, 1991-2008.

|  | A: Age & Sex<sup>a</sup> |  | B: Adding weight and<br>height<sup>a</sup> |  | C: Adding maternal<br>characteristics<sup>a</sup> |  |
| --- | --- | --- | --- | --- | --- | --- |
|  | Mean | 95% CI | Mean | 95% CI | Mean | 95% CI |
| Mean SBP: fixed effects |  |  |  |  |  |  |
| Intercept | 107.95 | 107.50, 108.40 | 107.93 | 107.52, 108.34 | 108.23 | 107.29, 109.16 |
| ≤12 years<sup>b</sup> | 1.71 | 1.58, 1.85 | -0.55 | -0.81, -0.30 | -0.56 | -0.83, -0.30 |
| >12 years<sup>b</sup> | 6.00 | 5.80, 6.21 | 3.83 | 3.51, 4.16 | 3.83 | 3.49, 4.18 |
| Female | -0.43 | -1.02, 0.19 | -0.40 | -0.96, 0.16 | -0.22 | -0.81, 0.36 |
| Female*≤12 years | 0.07 | -0.11, 0.25 | -0.15 | -0.33, 0.03 | -0.17 | -0.36, 0.01 |
| Female*>12 years | -1.99 | -2.28, -1.71 | -1.26 | -1.57, -0.95 | -1.25 | -1.58, -0.92 |
| log(Weight, kg) |  |  | 17.01 | 15.39, 18.64 | 16.56 | 14.84, 18.26 |
| Height (cm) |  |  | 0.04 | 0.00, 0.09 | 0.06 | 0.00, 0.11 |
| Maternal characteristics |  |  |  |  |  |  |
| Age at delivery (years) |  |  |  |  | 0.01 | -0.06, 0.09 |
| Parity |  |  |  |  | -0.17 | -0.50, 0.17 |
| Highest ed.: vocational<sup>c</sup> |  |  |  |  | -0.04 | -1.36, 1.30 |
| Highest ed.: O-level<sup>c</sup> |  |  |  |  | 0.05 | -0.95, 1.03 |
| Highest ed.: A-level<sup>c</sup> |  |  |  |  | -0.64 | -1.70, 0.40 |
| Highest ed.: Degree<sup>c</sup> |  |  |  |  | -1.39 | -2.50, -0.27 |
| Individual-level random effects for SBP |  |  |  |  |  |  |
| SD(mean BP)<sup>d</sup> | 6.12 | 5.88, 6.37 | 5.37 | 5.14, 5.60 | 5.39 | 5.16, 5.63 |
| SD(BP slope)<sup>d</sup> | 0.66 | 0.55, 0.75 | 0.64 | 0.54, 0.73 | 0.65 | 0.54, 0.74 |
| SD(log(BPV))<sup>d</sup> | 0.41 | 0.29, 0.51 | 0.39 | 0.26, 0.50 | 0.42 | 0.29, 0.53 |
| Cor(mean BP, BP slope)<sup>d</sup> | -0.09 | -0.20, 0.02 | -0.11 | -0.23, 0.00 | -0.09 | -0.21, 0.03 |
| Cor(mean BP, log(BPV))<sup>d</sup> | 0.52 | 0.36, 0.71 | 0.49 | 0.31, 0.72 | 0.50 | 0.33, 0.71 |
| Cor(BP slope, log(BPV))<sup>d</sup> | -0.06 | -0.34, 0.22 | -0.04 | -0.33, 0.26 | -0.05 | -0.34, 0.23 |
| BPV: fixed effects |  |  |  |  |  |  |
| Intercept | 3.22 | 3.12, 3.32 | 3.15 | 3.04, 3.25 | 3.13 | 3.02, 3.23 |
| Age (years)<sup>e</sup> | 15.60% | 12.87%, 18.39% | 11.28% | 5.80%, 16.98% | 10.76% | 5.05%, 16.77% |
| Female<sup>e</sup> | 16.66% | 4.53%, 30.40% | 17.44% | 4.66%, 31.59% | 19.83% | 5.99%, 35.46% |
| log(Weight, kg)<sup>f</sup> |  |  | 5.76% | 2.10%, 9.55% | 5.22% | 1.42%, 9.20% |
| Height (cm)<sup>e</sup> |  |  | -0.37% | -1.40%, 0.70% | -0.24% | -1.36%, 0.89% |
| Residual within-clinic SD for SBP | 5.62 | 5.54, 5.70 | 5.63 | 5.54, 5.71 | 5.61 | 5.53, 5.70 |
| Mean for log(LVMI): fixed effects |  |  |  |  |  |  |
| Intercept | 3.36 | 3.35, 3.37 | 3.33 | 3.31, 3.34 | 3.32 | 3.31, 3.33 |
| Age (years)<sup>e</sup> | -0.08% | -2.85%, 2.79% | -1.66% | -4.2%, 0.94% | -1.95% | -4.61%, 0.79% |
| Female<sup>e</sup> | -11.01% | -12.61%, -9.36% | -5.42% | -7.08%, -3.73% | -4.95% | -6.72%, -3.17% |
| Weight (kg)<sup>e</sup> |  |  | 0.69% | 0.62%, 0.76% | 0.70% | 0.63%, 0.78% |
| mean BP<sup>e,e</sup> | 0.85% | 0.44%, 1.44% | 0.57% | 0.09%, 1.49% | 0.59% | 0.12%, 1.37% |
| BP slope<sup>d,e</sup> | 4.36% | 0.88%, 8.61% | 3.60% | 0.00%, 8.35% | 3.56% | 0.01%, 7.92% |
| log(BPV)<sup>d,f</sup> | -0.61% | -2.05%, 0.40% | -0.83% | -3.08%, 0.29% | -0.78% | -2.54%, 0.22% |
| Residual SD for log(LVMI) | 0.20 | 0.19, 0.21 | 0.19 | 0.17, 0.19 | 0.19 | 0.18, 0.19 |

Abbreviations: BP: blood pressure; BPV: (within-individual) blood pressure variability; CI: credible interval; LVMI: left ventricular mass indexed to height.
<sup>a</sup> Sample sizes for Models A & B, N individuals: 1,986; N clinic visits: 10,556; N BP measurements: 19,360. Sample size for Model C, N individuals: 1,813; N clinic visits: 9,693; N BP measurements: 17,777.
<sup>b</sup> Linear spline terms, corresponding to change per year for age $\leq 12$ , and for age $>12$ , respectively.
<sup>c</sup> Reference category for mother's highest education qualification: CSE / None.
<sup>d</sup> mean BP denotes the individual-level, between-clinic, random effects for the intercept (at mean age 11.3 years) in the mean function for SBP; BP slope denotes the individual-level, between-clinic, random effects for age in the mean function for SBP; log(BPV) denotes the individual-level, between-clinic, random effects on the log-scale of the within-individual variance; i.e. these are the equivalent of $u_0$ , $u_1$ and $u_2$ in equation 3, respectively.
<sup>e</sup> estimates presented as percentage change in BPV / LVMI (on natural scale) per 1-unit increase in covariate; see Appendix 2 for regression coefficients.
<sup>f</sup> estimates presented as percentage change in BPV / LVMI (on natural scale) per 10% increase in covariate (on natural scale); see Appendix 2 for regression coefficients.

In model B, with weight and height now added as further covariates, mean SBP was predicted to increase with greater weight and height. With regard to BPV, this was estimated to be larger with greater log(weight): specifically, for a 10% increase in weight, BPV is predicted to increase by mean = 5.8% (95% CI: 2.1%, 9.6%) (SD(BP): mean = 2.6% (95% CI: 0.7%, 4.5%)). The estimated effect of height on log(BPV) was negative, with a substantial portion of the posterior containing positive values as well (BPV predicted to decrease by mean = −0.4% (95% CI: −1.4%, 0.7%) per 1cm increase in height). Otherwise, weight was predicted to have a positive association with LVMI.

Finally, in model C, maternal characteristics were added as covariates in the mean function for SBP (with a slight drop in sample size due to fewer observations for these variables). This predicted a modest positive association of mean SBP with mother’s age at delivery, lower mean SBP with higher parity, and with mean SBP generally predicted to be lower with more advanced highest educational qualifications. The estimated associations between the observed covariates of age, sex, height and weight and mean SBP were substantively similar whether maternal characteristics were included in the model (C) or not (B).

Focusing on Model C, this estimated a small negative correlation (*r* = −0.09 (95% CI: −0.21, 0.03)) between mean BP and BP slope: i.e. those with higher mean BP at 11.3 years were predicted to have a smaller increase in their mean BP across age. Otherwise, a large positive correlation (*r* = 0.50 (95% CI: 0.33, 0.71)) was estimated between mean BP and log(BPV): i.e. individuals with higher mean SBP at 11.3 years also tended to have more fluctuation in their SBP, whilst the correlation between BP slope and log(BPV) was estimated as negative, and small (*r* = −0.05 (95% CI: −0.34, 0.23)).

The SD of the BPV random effects on the log scale was estimated to be mean = 0.42 (95% CI: 0.29, 0.53). Figure 2 plots predicted within-individual SDs from this model, by sex, for the sex-specific mean value of the covariates within the within-individual variance function for each clinic. In addition, Figure 3 plots examples from this model of three individuals randomly-drawn from the 25 with the lowest, and 25 with the highest, estimated random effect for log(BPV) (with an added constant on both the x- and y-axis to preserve anonymity), illustrating instances of individual-level patterns of SBP measurements at each end of the observed range of estimated BPV.

**Figure 2.**
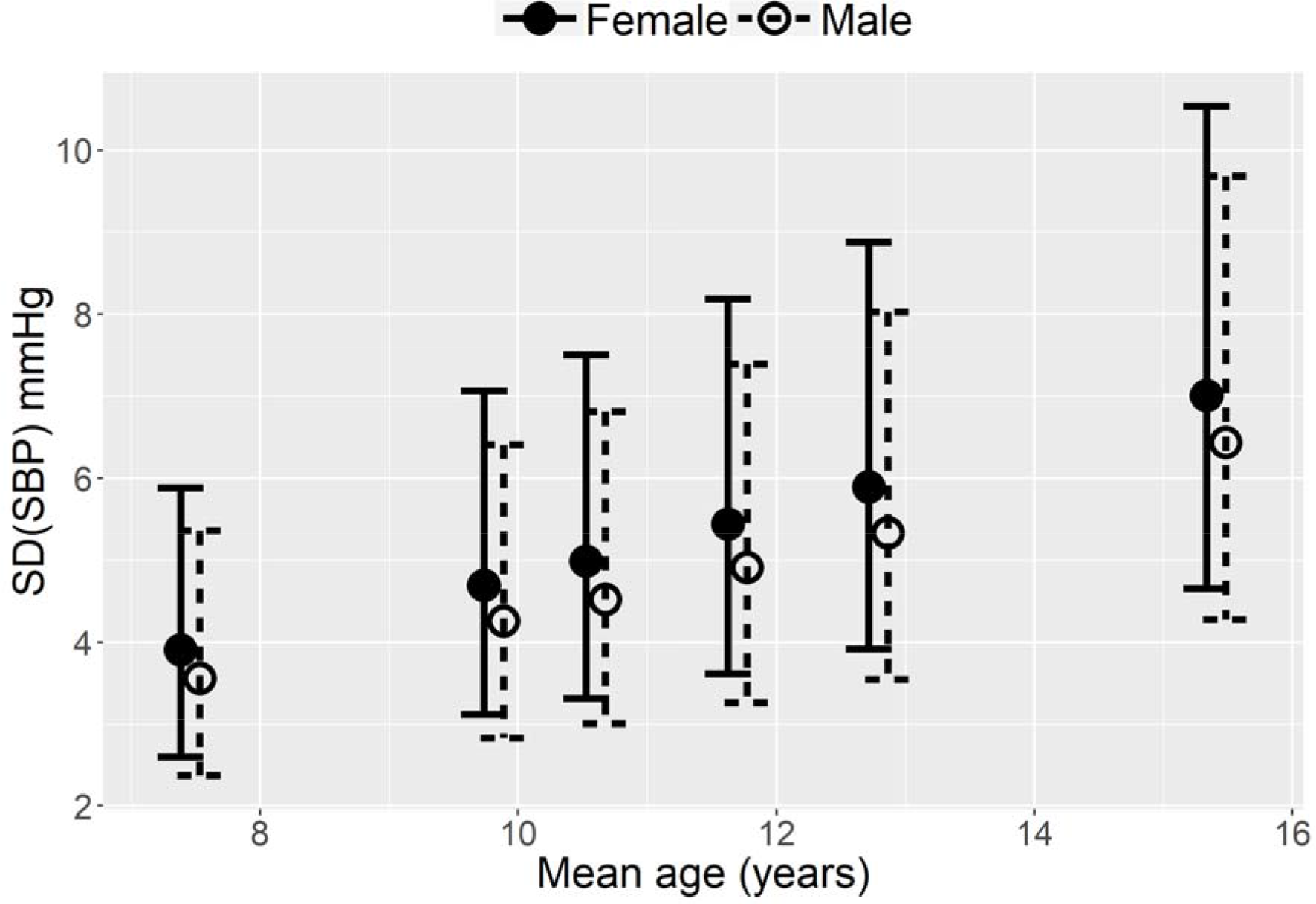
Predicted within-individual SD(SBP), by sex, for (sex-specific) mean values of age, height and weight at each research clinic. Mean with 95% confidence intervals. Avon Longitudinal Study of Parents And Children, SW England, 1991-2008. SBP, systolic blood pressure; SD, standard deviation.

**Figure 3.**
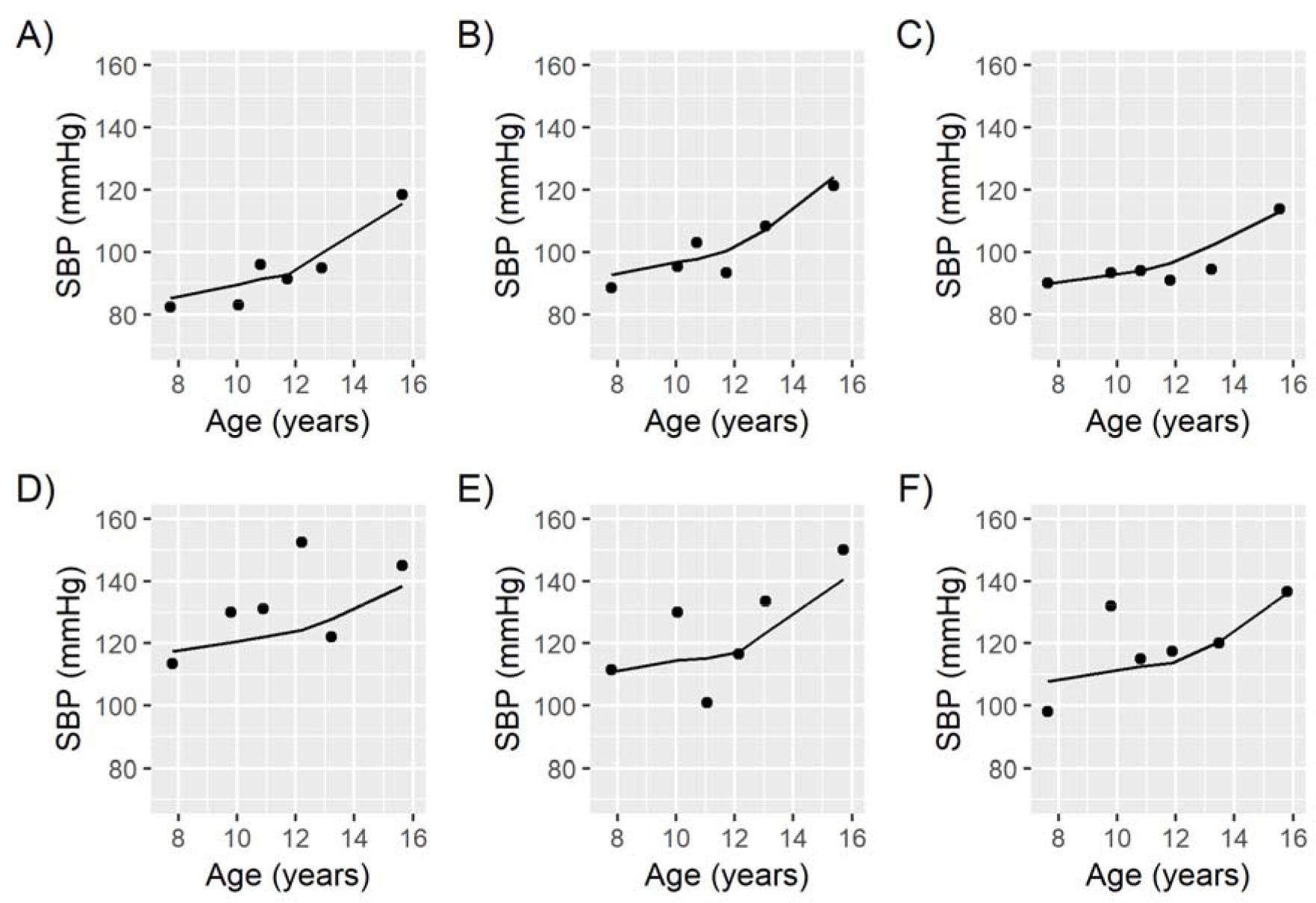
Observed data, and predictions, for three randomly-selected individuals with amongst the lowest (A-C), and three individuals with amongst the highest (D-F), random effect estimates for BPV. Note that a constant has been added to each axis to preserve anonymity. Avon Longitudinal Study of Parents And Children, SW England, 1991-2008. BPV, (within-individual) blood pressure variability; SBP, systolic blood pressure.

In the models in Table 2, all random effects have been included as predictors of the individual-level outcome, log(LVMI). Table 3 contrasts the estimates of these random effects from model C to those in which the effect of each set of random effects on log(LVMI) is tested without adjustment for the other random effects on log(LVMI) (but with the remaining structure of the model otherwise remaining the same; see Appendix 2 for full estimates). When mean BP is the only random effect included as a predictor of LVMI, higher mean BP is associated with higher LVMI. The same is true when the random slope and the log(BPV) random effects are alternately added as predictors of LVMI. With regard to the log(BPV) predictor, a 10% increase in BPV is predicted to increase LVMI by mean = 0.21% (95% CI: −0.23%, 0.69%) (or a 10% increase in SD(BP) is predicted to increase LVMI by mean = 0.42% (95% CI: −0.47%, 1.38%)). As Table 3 further indicates, when all three random effects are included as predictors of LVMI, then the estimated association between log(BPV) and LVMI becomes negative; specifically, a 10% increase in BPV is estimated to be associated with a decrease in LVMI by mean = −0.78% (95% CI: −2.54%, 0.22%) (or a 10% increase in SD(BP) is associated with a decrease in LVMI by mean = −1.56% (95% CI: −5.01%, 0.44%)). This model also estimated a positive association of mean BP with LVMI (a 1mmHg increase in BP at the sample mean age was predicted to increase LVMI by mean = 0.59% (95% CI: 0.12%, 1.37%)) and of change in mean BP with LVMI (a 1mmHg increase in the slope was predicted to increase LVMI by mean = 3.56% (95% CI: 0.01%, 7.92%)). Table 4 details mean LVMI for individuals grouped into tertiles of the random effect estimates for mean BP and log(BPV). It demonstrates that within strata of mean BP, higher values of BPV are generally associated with lower mean LVMI.

**Table 3.**
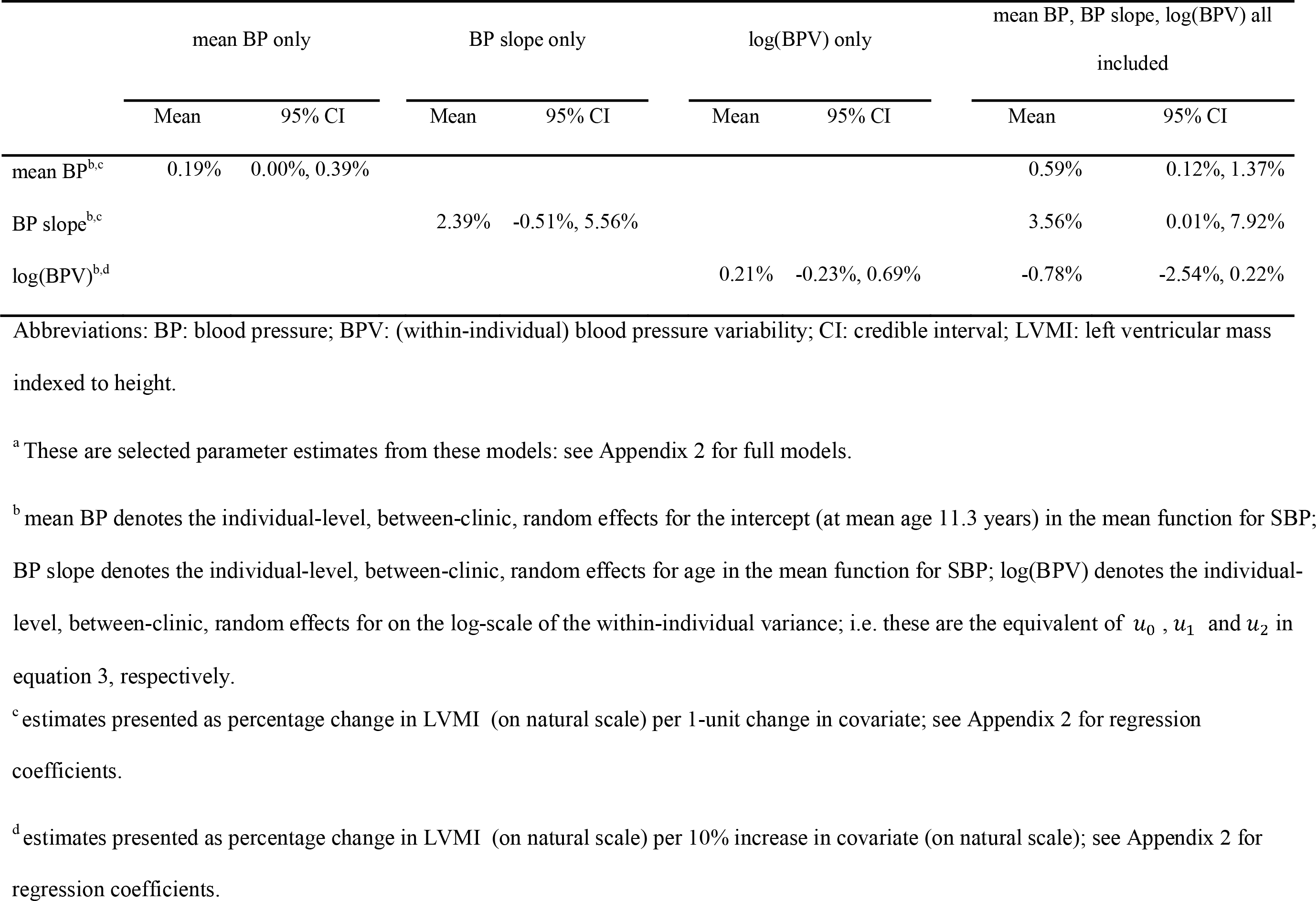
Posterior Parameter Estimates From Joint Models in Which Mean BP, BP Slope, and log(BPV) are Alternately Included as Exposures for log(LVMI).^a^ Avon Longitudinal Study of Parents And Children, SW England, 1991-2008.

|  | mean BP only |  | BP slope only |  | log(BPV) only |  | mean BP, BP slope, log(BPV) all included |  |
| --- | --- | --- | --- | --- | --- | --- | --- | --- |
|  | Mean | 95% CI | Mean | 95% CI | Mean | 95% CI | Mean | 95% CI |
| mean BP<sup>b,c</sup> | 0.19% | 0.00%, 0.39% |  |  |  |  | 0.59% | 0.12%, 1.37% |
| BP slope<sup>b,c</sup> |  |  | 2.39% | -0.51%, 5.56% |  |  | 3.56% | 0.01%, 7.92% |
| log(BPV)<sup>b,d</sup> |  |  |  |  | 0.21% | -0.23%, 0.69% | -0.78% | -2.54%, 0.22% |

Abbreviations: BP: blood pressure; BPV: (within-individual) blood pressure variability; CI: credible interval; LVMI: left ventricular mass indexed to height.
<sup>a</sup> These are selected parameter estimates from these models: see Appendix 2 for full models.
<sup>b</sup> mean BP denotes the individual-level, between-clinic, random effects for the intercept (at mean age 11.3 years) in the mean function for SBP; BP slope denotes the individual-level, between-clinic, random effects for age in the mean function for SBP; log(BPV) denotes the individual-level, between-clinic, random effects for on the log-scale of the within-individual variance; i.e. these are the equivalent of $u_0$ , $u_1$ and $u_2$ in equation 3, respectively.
<sup>c</sup> estimates presented as percentage change in LVMI (on natural scale) per 1-unit change in covariate; see Appendix 2 for regression coefficients.
<sup>d</sup> estimates presented as percentage change in LVMI (on natural scale) per 10% increase in covariate (on natural scale); see Appendix 2 for regression coefficients.

**Table 4.**
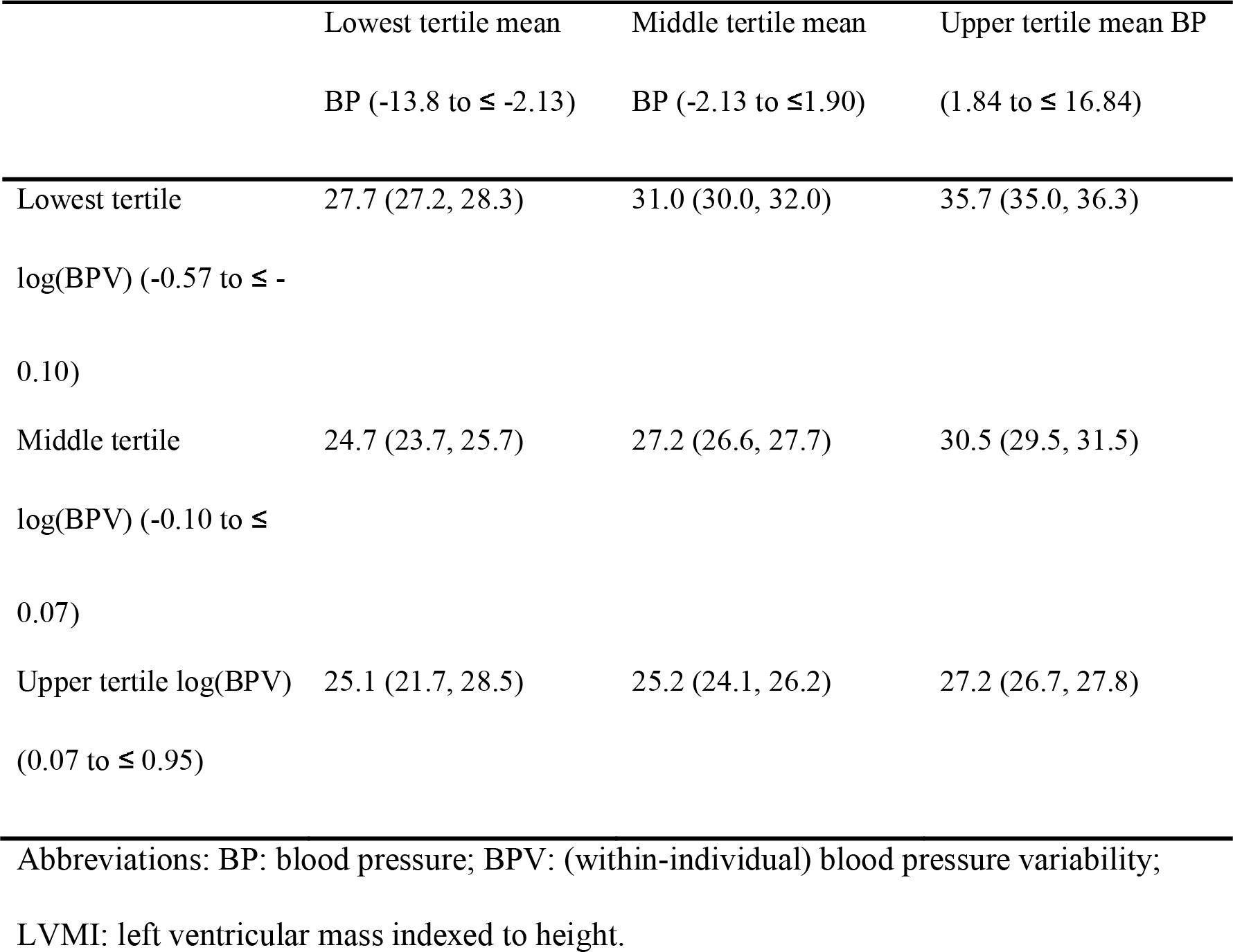
Mean LVMI (95% Confidence Interval), g/m^2.7^, for individuals grouped by tertiles of the random effect estimates for mean BP and log(BPV), Avon Longitudinal Study of Parents And Children, SW England, 1991-2008.

As Appendix 2 further discusses, a range of sensitivity analyses yielded substantively similar estimates to the models presented here.

## DISCUSSION

Using data from a prospective longitudinal birth cohort study cohort study based in the UK, we used a joint modelling approach to examine the factors associated with visit-to-visit BPV across childhood, and the association of both mean BP and BPV with an established biomarker of target organ damage, LVMI, in early adulthood. BPV was estimated to be larger – i.e. individuals’ SBP was estimated to fluctuate more greatly – at older ages, in females, and at greater bodyweights. BPV also had a large, positive correlation with mean BP (at the sample mean age of 11.3 years), but only a very small correlation with the rate of change in BP (slope) across age. In a model which did not adjust for the effect of mean BP and its slope on LVMI, BPV was estimated to be positively associated with LVMI. When mean BP and its slope were introduced as exposures of LVMI, however, the direction of association between BPV and LVMI changed to a negative one. These models further estimated a positive association of mean BP with LVMI and of change in mean BP with LVMI.

Whilst we are not aware of other studies in this age range which have reported on the effect of weight and height on BPV, the effect of age, sex and race have been examined. Investigating BPV in children aged 8-18 years in Massachusetts, Rosner *et al*. fitted variance components models to BP measurements taken within- and across-visits (up to a maximum of 16 visits over four years), and subsequently examined the effect of age, sex and BP level on these two variance components in three-way ANOVAs (16). They found “no meaningful effects” of these covariates on variability of SBP (the effect of BP level was significant, but judged to be small; correlation coefficient not reported). More recently, in an analysis of the Bogalusa Heart Study in the USA, Chen *et al*. measured visit-to-visit BPV in children aged 4-19 years by taking the SD of 4-8 measurements of clinic mean SBP from visits scheduled every 2-3 years (14). Males were estimated to have significantly higher SD(SBP) than females, and SD(SBP) was estimated to be significantly greater for blacks than for whites. Our supplementary analyses (Appendix 2) also indicated that females were estimated to have lower BPV than males, but not once the effect of time-varying covariates were adjusted for. In addition, and in keeping with our findings, Chen *et al*. found a positive correlation (*r* = 0.15) between mean childhood BP level and SD(SBP) (14). Such a positive correlation has been characterised in studies of adults too (8, 33).

Whilst several studies have found a positive relationship between adult BPV, independent of mean BP, and later cardiovascular events such as stroke (8) and all-cause mortality (33), evidence for the relationship between BPV and target organ damage (as indicated by left ventricular hypertrophy, for example) is more equivocal (34, 35). In a recent review, Dolan & O’Brien described contrasting results with regard to the association of visit-to-visit BPV and cardiovascular outcomes, suggesting differences between (adult) study populations, such as underlying cardiovascular risk, may be an important determinant of such heterogenous findings (7). With regard to the relationship between childhood BPV and adult biomarkers, Chen *et al*. found childhood visit-to-visit SD(SBP) to be significantly associated with adult hypertension (14). This remained the case after adjusting for mean childhood SBP, although elevated childhood SBP levels were found to be more predictive of adult hypertension than childhood BPV. We have previously demonstrated that in 17 year-olds, higher body mass index (BMI) is causally related to higher LVMI, suggesting that there is meaningful variation in cardiac structure measures in early adulthood that is likely to track across life and relate to later life cardiovascular health (36). Nevertheless, in the current study the positive association between childhood BPV and LVMI in early adulthood did not persist once childhood mean BP had been adjusted for. In fact, within strata of mean childhood SBP, higher BPV was associated with smaller values of LVMI. Whether it is appropriate to adjust for mean BP when assessing the association of BPV with LVMI somewhat depends on whether one has a causal model in mind which conceptualises mean BP as causing both BPV and LVMI (in which case adjustment for mean BP as a confounder might be appropriate), or whether no causal relationship is proposed between mean BP and BPV (i.e. other, unknown, factors cause mean BP and BPV, and each in turn has a causal relationship with later LVMI), in which case it may not be appropriate to adjust for mean BP. If the purpose is prediction, on the other hand, then by simultaneously estimating the effect of mean BP, its slope, and BPV on the later outcome, we allow for a more complete appraisal of the association of repeatedly-measured BP with LVMI. Note that whether mean BP is adjusted for or not, the association of BPV with LVMI was estimated to be small, with appreciable uncertainty as to whether it was non-zero, or indeed of the opposite sign.

The results also indicated that mean BP (at the sample mean age), and change in mean BP, were both positively associated with LVMI in early adulthood. Analyses of the Georgia Stress and Heart Study and the Bogalusa Heart Study have also found mean BP in childhood (37), and its slope from childhood to young adulthood (38) and in adolescence (39), to be positively associated with left ventricular hypertrophy in adulthood.

If we assume that selection into our analysis only depends on measured variables such as sex of child, education and age of mother, then their inclusion as covariates will result in unbiased models. If, having conditioned on such covariates, inclusion in our models is related to the exposure (SBP) and/or outcome (LVMI), then there will be bias. *A priori*, this seems relatively unlikely, however: e.g. such biomarkers are not readily-observable in a manner which would typically influence one’s participation in a study. Whilst there were fewer observations for the maternal characteristics (*cf*. child-based covariates) included in some models, the proportion of missing data was relatively modest (8-9%), and estimates for parameters common to each model were substantively similar.

The general class of model we have used could be employed to examine within-person variability in any repeatedly-measured exposure, relating it to later individual-level outcomes of interest within a joint model: e.g. the association between glycaemic variability and mortality (40), variability in gait and risk of falling in older adults (41), and variation in prostate specific antigen and prostate volume (42). We have focused on within-individual variability, but interest may lie in characterising within-clinic (inter-individual) variability instead. In addition, the models can be readily adapted for binary variables, time-to-event data (3), etc. (with additional adjustments made as appropriate to accommodate e.g. autocorrelation in the case of intensive longitudinal measurements such as ambulatory BP).

We have supplied example code to demonstrate how these models can be fitted using a Bayesian framework (using Stan (30) and BUGS (43): see Appendix 3, which also includes discussion of how many repeated measures are needed to estimate these models. Endeavours to fit such models are being actively extended to other software packages too (44, 45)). Whilst fitting these models involves greater computational burden and complexity compared to a two-stage approach, the latter is known to introduce bias (3). Joint models have been recently employed to examine within-individual variability (3). We further extend these by investigating the association of time-varying covariates with within-individual variability, allowing us to examine residual BPV unexplained by known factors, and also by distinguishing between-clinic from within-clinic variability. As such, the joint modelling approach we have introduced offers a very flexible method of exploring the factors associated with within-individual variability.

## Data Availability

Data are available following relevant procedures for data access from Avon Longitudinal Study of Parents and Children (ALSPAC, http://www.bristol.ac.uk/alspac/researchers/access/); computing code to fit models to simulated data is provided in Appendix 3 of the Supplementary Materials.

## ACKNOWLEDGEMENTS

We are extremely grateful to all the families who took part in this study, the midwives for their help in recruiting them, and the whole ALSPAC team, which includes interviewers, computer and laboratory technicians, clerical workers, research scientists, volunteers, managers, receptionists and nurses.

## ABBREVIATIONS

BMI: body mass index
BPV: (within-individual) blood pressure variability
BP: blood pressure
CI: credible interval
CVD: cardiovascular disease
LVMI: left ventricular mass indexed to height
SBP: systolic blood pressure
SD: standard deviation

## REFERENCES

1. Tielemans S, Geleijnse JM, Menotti A, et al. Ten-Year Blood Pressure Trajectories, Cardiovascular Mortality, and Life Years-Lost in 2 Extinction Cohorts: the Minnesota Business and Professional Men Study and the Zutphen Study. J Am Heart Assoc 2015;4(3).

2. Elliott MR, Sammel MD, Faul J. Associations between variability of risk factors and health outcomes in longitudinal studies. Statistics in Medicine 2012;31(23):2745–56.

3. Barrett JK, Huille R, Parker RMA, et al. Estimating the association between blood pressure variability and cardiovascular disease: An application using the ARIC Study. Statistics in Medicine 2019;38(10):1855–68.

4. Tu YK, Tilling K, Sterne JAC, et al. A critical evaluation of statistical approaches to examining the role of growth trajectories in the developmental origins of health and disease. International Journal of Epidemiology 2013;42(5):1327–39.

5. Sayers A, Heron J, Smith A, et al. Joint modelling compared with two stage methods for analysing longitudinal data and prospective outcomes: A simulation study of childhood growth and BP. Statistical Methods in Medical Research 2017;26(1):437–52.

6. Sweeting MJ, Thompson SG. Joint modelling of longitudinal and time-to-event data with application to predicting abdominal aortic aneurysm growth and rupture. Biometrical Journal 2011;53(5):750–63.

7. Dolan E, O’Brien E. Is It Daily, Monthly, or Yearly Blood Pressure Variability that Enhances Cardiovascular Risk? Current Cardiology Reports 2015;17(11).

8. Rothwell PM, Howard SC, Dolan E, et al. Prognostic significance of visit-to-visit variability, maximum systolic blood pressure, and episodic hypertension. Lancet 2010;375(9718):895–905.

9. Berenson GS, Wattigney WA, Tracy RE, et al. Atherosclerosis of the aorta and coronary-arteries and cardiovascular risk-factors in persons aged 6 to 30 years and studied at necropsy (the Bogalusa Heart Study). American Journal of Cardiology 1992;70(9):851–8.

10. Berenson GS, Bogalusa Heart Study Res G. Childhood risk factors predict adult risk associated with subclinical cardiovascular disease: The Bogalusa Heart Study. American Journal of Cardiology 2002;90(10C):3L–7L.

11. Laird WP, Fixler DE. Left-ventricular hypertrophy in adolescents with elevated blood-pressure - assessment by chest roentgenography, electrocardiography, and echocardiography. Pediatrics 1981;67(2):255–9.

12. Malcolm DD, Burns TL, Mahoney LT, et al. Factors affecting left-ventricular mass in childhood - the Muscatine Study. Pediatrics 1993;92(5):703–9.

13. Daniels SR, Kimball TR, Morrison JA, et al. Effect of lean body-mass, fat mass, blood-pressure, and sexual-maturation on left-ventricular mass in children and adolescents - statistical, biological, and clinical-significance. Circulation 1995;92(11):3249–54.

14. Chen W, Srinivasan SR, Ruan LT, et al. Adult Hypertension Is Associated With Blood Pressure Variability in Childhood in Blacks and Whites: The Bogalusa Heart Study. American Journal of Hypertension 2011;24(1):77–82.

15. Whincup PH, Nightingale CM, Owen CG, et al. Ethnic Differences in Carotid Intima-Media Thickness Between UK Children of Black African-Caribbean and White European Origin. Stroke 2012;43(7):1747–54.

16. Rosner B, Cook NR, Evans DA, et al. Reproducibility and predictive values of routine blood-pressure measurements in children - comparison with adult values and implications for screening-children for elevated blood-pressure. American Journal of Epidemiology 1987;126(6):1115–25.

17. Boyd A, Golding J, Macleod J, et al. Cohort Profile: The ‘Children of the 90s’-the index offspring of the Avon Longitudinal Study of Parents and Children. International Journal of Epidemiology 2013;42(1):111–27.

18. Fraser A, Macdonald-Wallis C, Tilling K, et al. Cohort Profile: The Avon Longitudinal Study of Parents and Children: ALSPAC mothers cohort. International Journal of Epidemiology 2013;42(1):97–110.

19. Northstone K, Lewcock M, Groom A, et al. The Avon Longitudinal Study of Parents and Children (ALSPAC): an updated on the enrolled sample of index children in 2019. Wellcome Open Research 2019 2019;4:51.

20. Timpka S, Hughes AD, Chaturvedi N, et al. Birth weight and cardiac function assessed by echocardiography in adolescence: Avon Longitudinal Study of Parents and Children. Ultrasound in Obstetrics & Gynecology 2019;54(2):225–31.

21. Timpka S, Macdonald-Wallis C, Hughes AD, et al. Hypertensive Disorders of Pregnancy and Offspring Cardiac Structure and Function in Adolescence. J Am Heart Assoc 2016;5(11).

22. Hedeker D, Mermelstein RJ, Demirtas H. An application of a mixed-effects location scale model for analysis of ecological momentary assessment (EMA) data. Biometrics 2008;64(2):627–34.

23. Hedeker D, Mermelstein RJ, Demirtas H. Modeling between-subject and within-subject variances in ecological momentary assessment data using mixed-effects location scale models. Statistics in Medicine 2012;31(27):3328–36.

24. Goldstein H, Leckie G, Charlton C, et al. Multilevel growth curve models that incorporate a random coefficient model for the level 1 variance function. Stat Methods Med Res 2017:962280217706728.

25. Rast P, Hofer SM, Sparks C. Modeling Individual Differences in Within-Person Variation of Negative and Positive Affect in a Mixed Effects Location Scale Model Using BUGS/JAGS. Multivariate Behavioral Research 2012;47(2):177–200.

26. Staley JR, Bradley J, Silverwood RJ, et al. Associations of Blood Pressure in Pregnancy With Offspring Blood Pressure Trajectories During Childhood and Adolescence: Findings From a Prospective Study. J Am Heart Assoc 2015;4(5):12.

27. O’Keeffe LM, Simpkin AJ, Tilling K, et al. Sex-specific trajectories of measures of cardiovascular health during childhood and adolescence: A prospective cohort study. Atherosclerosis 2018;278:190–6.

28. Howe LD, Tilling K, Matijasevich A, et al. Linear spline multilevel models for summarising childhood growth trajectories: A guide to their application using examples from five birth cohorts. Statistical Methods in Medical Research 2016;25(5):1854–74.

29. Stan Development Team. Stan User’s Guide. 2018.

30. Carpenter B, Gelman A, Hoffman MD, et al. Stan: A Probabilistic Programming Language. Journal of Statistical Software 2017;76(1):1–29.

31. R Core Team. R: A Language and Environment for Statistical Computing. Vienna, Austria: R Foundation for Statistical Computing, 2018.

32. Stan Development Team. RStan: the R interface to Stan. 2018.

33. Muntner P, Shimbo D, Tonelli M, et al. The Relationship Between Visit-to-Visit Variability in Systolic Blood Pressure and All-Cause Mortality in the General Population Findings From NHANES III, 1988 to 1994. Hypertension 2011;57(2):160–6.

34. Vishram JKK, Dahlof B, Devereux RB, et al. Blood pressure variability predicts cardiovascular events independently of traditional cardiovascular risk factors and target organ damage: a LIFE substudy. Journal of Hypertension 2015;33(12):2422–30.

35. Veloudi P, Blizzard CL, Head GA, et al. Blood Pressure Variability and Prediction of Target Organ Damage in Patients With Uncomplicated Hypertension. American Journal of Hypertension 2016;29(9):1046–54.

36. Wade KH, Chiesa ST, Hughes AD, et al. Assessing the Causal Role of Body Mass Index on Cardiovascular Health in Young Adults Mendelian Randomization and Recall-by-Genotype Analyses. Circulation 2018;138(20):2187–201.

37. Toprak A, Wang HW, Chen W, et al. Relation of childhood risk factors to left ventricular hypertrophy (eccentric or concentric) in relatively young adulthood (from the Bogalusa Heart Study). American Journal of Cardiology 2008;101(11):1621–5.

38. Hao G, Wang XL, Treiber FA, et al. Blood Pressure Trajectories From Childhood to Young Adulthood Associated With Cardiovascular Risk Results From the 23-Year Longitudinal Georgia Stress and Heart Study. Hypertension 2017;69(3):435]+.

39. Zhang T, Li SX, Bazzano L, et al. Trajectories of Childhood Blood Pressure and Adult Left Ventricular Hypertrophy: The Bogalusa Heart Study. Hypertension 2018;72(1):93–101.

40. Hirakawa Y, Arima H, Zoungas S, et al. Impact of Visit-to-Visit Glycemic Variability on the Risks of Macrovascular and Microvascular Events and All-Cause Mortality in Type 2 Diabetes: The ADVANCE Trial. Diabetes care 2014.

41. Hausdorff JM, Rios DA, Edelberg HK. Gait variability and fall risk in community-living older adults: A 1-year prospective study. Archives of Physical Medicine and Rehabilitation 2001;82(8):1050–6.

42. Nichols JH, Loeb S, Metter EJ, et al. The relationship between prostate volume and prostate-specific antigen variability: data from the Baltimore Longitudinal Study of Aging and the Johns Hopkins Active Surveillance Program. BJU international 2012;109(9):1304–8.

43. Lunn DJ, Thomas A, Best N, et al. WinBUGS — a Bayesian modelling framework: concepts, structure, and extensibility. Statistics and Computing 2000;10:325–37.

44. Crowther MJ. merlin - a unified modelling framework for data analysis and methods development in Stata. arXivorg 2018;1806.01615v1.

45. Hedeker D, Dunton GF. MIXWILD User’s Guide. University of Chicago & University of Southern California; 2018.

